# Characterizing gun violence by time, day of the week, and holidays in six US cities 2015-2021

**DOI:** 10.1101/2022.12.28.22283963

**Authors:** Elizabeth B. Klerman, Mahmoud Affouf, Rebecca Robbins, Jay Iyer, Peter T. Masiakos, Chana A. Sacks

## Abstract

Developing interventions to prevent firearm-related violence and to address its consequences requires an improved understanding of when these violent events are most likely to occur. We explored gunshot events by time of day, day of week, and holiday/non-holiday in six major US cities. We utilized publicly available police department datasets that report the date and time of day of gunshot events from six major US cities. In these cities, shootings occurred most often at night and on holidays and weekends, with significant interaction effects. Primary prevention efforts should consider this differential risk.

## Background

The United States of America has a death rate due to gun violence higher than that of other high-income countries^1^; firearm related injuries are now the leading cause of death in children and adolescents in the USA^2^. Some violent behaviors occur more often at night (reviewed in ^3^). Developing interventions to prevent firearm-related violence and to address its consequences requires an improved understanding of when these violent events are most likely to occur. We explored gunshot events by time of day, day of week, and holiday/non-holiday in six major US cities.

## Methods

We utilized publicly available police department datasets (listed in **Reference** section) that report the date and time of day of gunshot events in Baltimore MD, Boston MA, Washington DC, New York City (NYC) NY, Philadelphia PA, and Portland OR. Datasets from Baltimore, NYC, Philadelphia, and Portland included information on when gunshots events resulted in a victim; the dataset from Washington DC reported information on gunshots detected from a technology surveillance system (Shotspotter), regardless of whether there was a victim; the Boston datasets reported gunshot events both with and without victims. Data from Baltimore, Boston, Washington DC, NYC and Philadelphia were from Jan 1, 2015 -Dec 31, 2021 and from Portland were from Jan 1, 2019-Dec 31, 2021. All cities reported data by date; all cities except Portland reported data by time (HH:MM); Portland reported data in 2-hour bins starting at midnight. Therefore, we summed the counts of gunshot events into 2-hour bins for all cities.

Holidays were defined as days in which government offices were closed and were identified according to federal, state and city calendars. Nighttime was defined as 18:00-05:59 and daytime as 06:00-17:59 based on visual inspection of the data from all cities. Weekend was defined as Friday 18:00-Sunday 17:59, Weekday as Sunday 18:00-Friday 17:59, and Monday holidays as Sunday 18:00-Monday 17:59.

For each city, we summed the number of gunshot events by time of day, day of week, and holiday/non-holiday. We analyzed by nighttime vs. daytime overall and within day of week, weekday holidays vs non-holidays, and weekend vs. weekday and their interactions. We computed ratios of the number of gunshot events of nighttime/daytime, and for nighttime and daytime separately for weekends, for weekday holiday and non-holiday, and for Monday holiday and non-holiday. We adjusted for the number of days in each group for these ratios.

## Results (Figure 1, Table 1)

The ratio of nighttime/daytime gunshot events ranged from 1.7 - 4.5.

**Figure 1:**
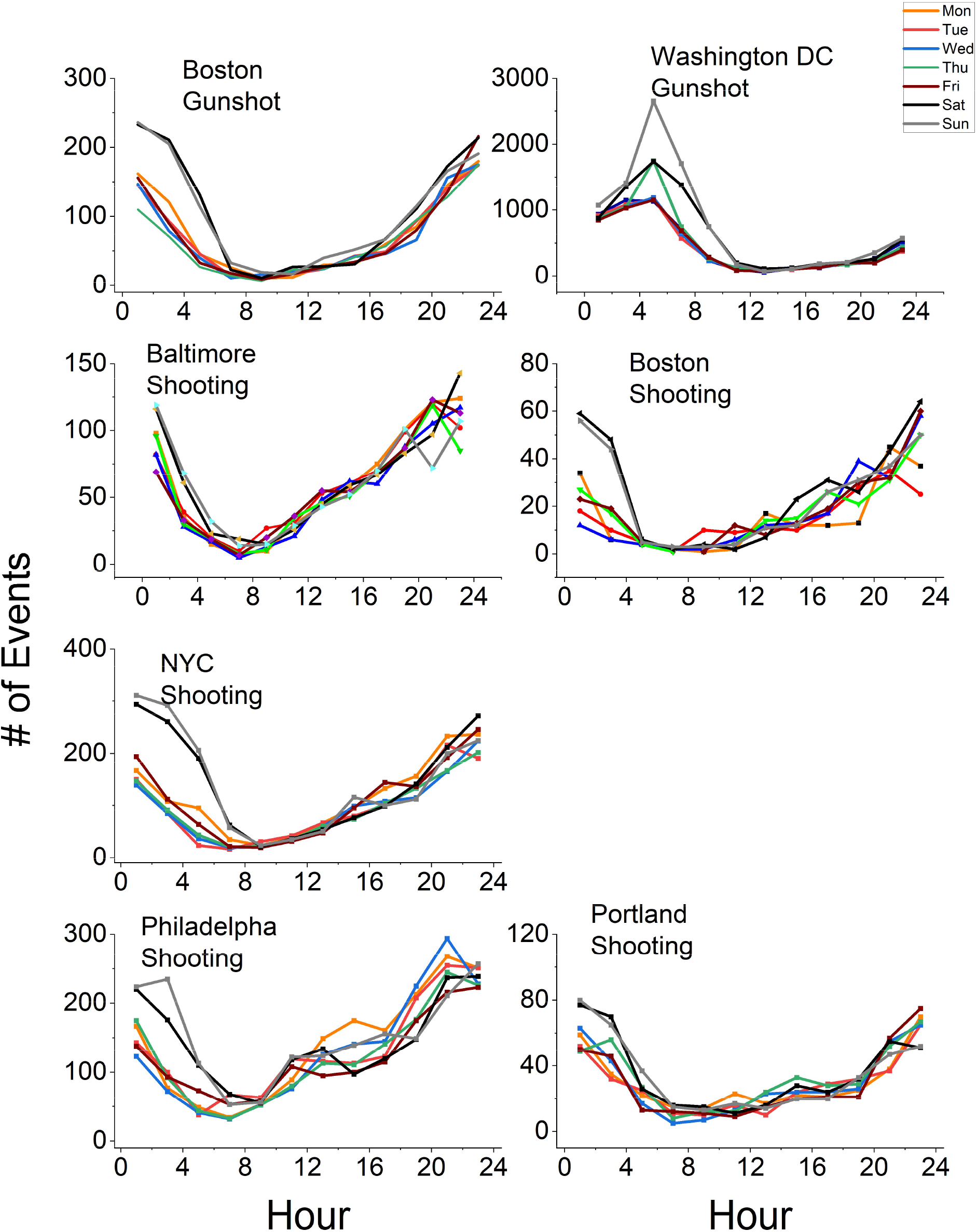
Number of gunshots or shootings for each city by time of calendar day and day of week.

**Table 1:** Relative rations of gunshots or shootings for each city and event type

|  | Baltimore | Boston | Boston | Washington,<br>DC | NYC | Philadelphia | Portland |
| --- | --- | --- | --- | --- | --- | --- | --- |
|  | Shooting | Gunshot | Shooting | Gunshot | Shooting | Shooting | Shooting |
| Night/Day | 2.1 | 4.5 | 3.0 | 2.6 | 2.8 | 1.7 | 2.7 |
| **Nighttime** |  |  |  |  |  |  |  |
| Weekday<br>holiday/non-holiday | 0.9 | 1.3 | 1.7 | 1.7 | 1.5 | 1.1 | 1.3 |
| Monday<br>holiday/non-holiday | 1.0 | 1.2 | 1.3 | 1.1 | 1.6 | 0.9 | 0.9 |
| Weekend/<br>Weekday<br>non-holiday | 1.2 | 1.5 | 1.7 | 1.3 | 1.7 | 1.2 | 1.3 |
| **Daytime** |  |  |  |  |  |  |  |
| Weekday<br>holiday/non-holiday | 1.3 | 1.2 | 1.0 | 2.4 | 1.0 | 0.9 | 1.0 |
| Monday<br>holiday/non-holiday | 1.3 | 1.4 | 0.8 | 1.5 | 1.1 | 0.7 | 0.9 |
| Weekend/<br>Weekday<br>non-holiday | 1.0 | 1.3 | 1.2 | 2.0 | 1.1 | 1.1 | 1.0 |

Nighttime is defined as hours 18:00-0559; daytime as 06:00-17:59.
Weekends and holidays are defined as beginning at 18:00 the calendar day before and ending at 17:59 that calendar day.

The ratios of gunshot events on weekdays that were holidays/non-holidays during daytime ranged from 0.9-2.4; during nighttime, these ratios ranged from 0.9-1.7. The ratios of gunshot events on Mondays that were holidays/non-holidays during daytime ranged from 0.7-1.5; during nighttime, these ratios ranged from 0.9-1.6.

The relative ratio of gunshot events on weekend/weekday non-holidays during daytime ranged from 1.0-2.0 and during nighttime ranged from 1.2-1.7.

## Discussion

In these six major US cities, shootings occurred most often at night and on holiday and weekend days, with significant interaction effects.

The results suggest that gunshot events are affected by standard work or school schedules, since there were fewer gunshot events during the day and on non-holidays or weekends.

Primary prevention efforts should consider this differential risk. Beyond the impact on individuals, the sounds of gun shots and the lights and sirens of emergency vehicles are likely to disrupt the neighborhoods in which this violence occurs. Because sufficient sleep with minimal interruption during the night supports daytime cognition and emotional regulation and is vital for mental health, learning, and physical health, these night-time predominant patterns of gun violence may be an important factor in understanding the community-level consequences of firearm-related violence.

## Data Availability

Data are available online at the links provided in the Data Availability Links section.

https://data.baltimorecity.gov/datasets/baltimore::part1-crime-data/explore?showTable=true

https://data.boston.gov/organization/boston-police-department-org

https://data.boston.gov/dataset/shootings

https://mpdc.dc.gov/node/1310761

https://data.cityofnewyork.us/Public-Safety/NYPD-Shooting-Incident-Data-Historic-/833y-fsy8

https://www.opendataphilly.org/dataset/shooting-victims

https://public.tableau.com/app/profile/portlandpolicebureau/viz/PortlandShootingIncidentStatistics/ShootingIncidentStatistics

## Data sources for gunshots and/or shooting for each city

| City | Data Type | Link(s) to Data |
| --- | --- | --- |
| Baltimore, MD | Gunshots | https://data.baltimorecity.gov/datasets/baltimore::part1-crime-data/explore?showTable=true |
| Boston, MA | Gunshots | https://data.boston.gov/organization/boston-police-department-org |
| Boston, MA | Shooting | https://data.boston.gov/dataset/shootings |
| Washington, DC | Gunshots | https://mpdc.dc.gov/node/1310761 |
| New York City, NY | Shooting | https://data.cityofnewyork.us/Public-Safety/NYPD-Shooting-Incident-Data-Historic-/833y-fsy8 |
| Philadelphia, PA | Shooting | https://www.opendataphilly.org/dataset/shooting-victims |
| Portland, OR | Shooting | https://public.tableau.com/app/profile/portlandpolicebureau/viz/PortlandShootingIncidentStatistics/ShootingIncidentStatistics |

## Support

EBK: NIH R01-NS099055, U01-NS114001, U54-AG062322, R21-DA052861, R21-DA052861, R01-NS114526-02S1, R01-107064; DoD W81XWH201076; and Leducq Foundation for Cardiovascular Research

MA: (none)

RR: NIH K01-HL150339

JI: (none)

PTM: (none)

CAS: (none)

## COI (2021-present)

EBK: *consulting*: American Academy of Sleep Medicine Foundation, Circadian Therapeutics, National Sleep Foundation, Sleep Research Society Foundation, Yale University Press. *Travel support*: European Biological Rhythms Society. *Other*: partner owns Chronsulting.

MA:(none)

RR: *consulting:* Sonesta Hotels, Castle Hot Springs, Oura Ring, Savoir Beds, byNacht GmbH.

JI: (none)

PTM: (none)

CAS: (none)

## Notes

### Competing Interest Statement

MA, JI, PTM, CAS: none
EBK: consulting: American Academy of Sleep Medicine Foundation, Circadian Therapeutics, National Sleep Foundation, Sleep Research Society Foundation, Yale University Press. Travel support: European Biological Rhythms Society. Other: partner owns Chronsulting.
RR: consulting: Sonesta Hotels, Castle Hot Springs, Oura Ring, Savoir Beds, byNacht GmbH.
MA

### Funding Statement

MA, JI, PTM, CAS: None
EBK: NIH R01-NS099055, U01-NS114001, U54-AG062322, R21-DA052861, R21-DA052861, R01-NS114526-02S1, R01-107064; DoD W81XWH201076; and Leducq Foundation for Cardiovascular Research
RR: NIH K01-HL150339

### Author Declarations

Source data were openly available before the initiation of the study. Links are: https://data.baltimorecity.gov/datasets/baltimore::part1-crime-data/explore?showTable=true https://data.boston.gov/organization/boston-police-department-org https://data.boston.gov/dataset/shootings https://mpdc.dc.gov/node/1310761 https://data.cityofnewyork.us/Public-Safety/NYPD-Shooting-Incident-Data-Historic-/833y-fsy8 https://www.opendataphilly.org/dataset/shooting-victims https://public.tableau.com/app/profile/portlandpolicebureau/viz/PortlandShootingIncidentStatistics/ShootingIncidentStatistics

## References

1. Grinshteyn, E. & Hemenway, D. Violent Death Rates: The US Compared with Other High-income OECD Countries, 2010. The American Journal of Medicine 129, 266–273 (2016).

2. Goldstick, J. E., Cunningham, R. M. & Carter, P. M. Current Causes of Death in Children and Adolescents in the United States. N Engl J Med 386, 1955–1956 (2022).

3. Tubbs, A. S., Fernandez, F.-X., Grandner, M. A., Perlis, M. L. & Klerman, E. B. The Mind After Midnight: Nocturnal Wakefulness, Behavioral Dysregulation, and Psychopathology. Front. Netw. Physiol. 1, 830338 (2022).

